# HormoneBayes: a novel Bayesian framework for the analysis of pulsatile hormone dynamics

**DOI:** 10.1101/2022.03.14.22272000

**Authors:** Margaritis Voliotis, Ali Abbara, Julia K Prague, Johannes D. Veldhuis, Waljit S Dhillo, Krasimira Tsaneva-Atanasova

## Abstract

The hypothalamus is the central regulator of reproductive hormone secretion. Pulsatile secretion of gonadotropin releasing hormone (GnRH) is fundamental to physiological stimulation of the pituitary gland to release luteinizing hormone (LH) and follicle stimulating hormone (FSH). Furthermore, GnRH pulsatility is altered in common reproductive disorders such as polycystic ovary syndrome (PCOS) and hypothalamic amenorrhea (HA). LH is measured routinely in clinical practice using an automated chemiluminescent immunoassay method and is the gold standard surrogate marker of GnRH. LH can be measured at frequent intervals (e.g., 10 minutely) to assess GnRH/LH pulsatility. However, this is rarely done in clinical practice because it is resource intensive, and there is no user-friendly and accessible method for computational analysis of the LH data available to clinicians. Here we present *hormoneBayes*, a novel open-access Bayesian framework that can be easily applied to reliably analyze serial LH measurements to assess LH pulsatility. The framework utilizes parsimonious models to simulate hypothalamic signals that drive LH dynamics, together with state-of-the-art (sequential) Monte-Carlo methods to infer key parameters and latent hypothalamic dynamics. We show that this method provides estimates for key pulse parameters including inter-pulse interval, secretion and clearance rates and identifies LH pulses in line with the current gold-standard deconvolution method. We show that these parameters can distinguish LH pulsatility in different clinical contexts including in reproductive health and disease in men and women (e.g., healthy men, healthy women before and after menopause, women with HA or PCOS). A further advantage of *hormoneBayes* is that our mathematical approach provides a quantified estimation of uncertainty. Our framework will complement methods enabling real-time *in-vivo* hormone monitoring and therefore has the potential to assist translation of personalized, data-driven, clinical care of patients presenting with conditions of reproductive hormone dysfunction.

## Introduction

Pulsatile hormone dynamics are ubiquitous and play a crucial role in the regulation of many bodily functions related to metabolism, stress, and fertility ^1,2^. Hormones are typically secreted in both a basal manner to maintain steady state levels, as well as with superimposed interspersed transient bursts (pulses) ^3^. It is now established that the pulsatile nature of hormonal secretion affects their interaction with receptors and downstream effector action ^4-6^.

With regards to fertility, the hypothalamus is the central regulator of the reproductive endocrine axis. Notably, gonadotropin releasing hormone (GnRH) is secreted in a pulsatile manner, and seminal studies have demonstrated that this pulsatility is fundamental for its action to stimulate GnRH receptors on pituitary gonadotropes ^5^. Moreover, disturbances in GnRH pulsatility are observed in common reproductive disorders including polycystic ovary syndrome (PCOS) in which GnRH pulsatility is increased ^7^, and hypothalamic amenorrhea (HA) in which GnRH pulsatility is reduced ^8^. However, despite this disparate alteration in GnRH pulsatility, differentiation of these two common reproductive disorders, which may both present similarly with menstrual disturbance, can be challenging ^9^. LH is measured routinely in clinical practice using an automated chemiluminescent immunoassay method and is the gold standard surrogate marker of GnRH. Furthermore, LH can be measured at frequent intervals (eg 10minutely) to assess GnRH/LH pulsatility, and accurate assessment of LH pulsatility could help facilitate diagnosis and treatment of patients presenting with reproductive endocrine disorders ^9^. However, this is rarely done in clinical practice because it is resource-intensive, inconvenient for patients, and there is a lack of user-friendly methods for computational analysis of the LH data available to clinicians.

Analysis of hormone pulsatility is a challenging computational problem, primarily due the stochastic nature of hormone dynamics and the consequent pulse-to-pulse variability, but also due to extrinsic factors (such as measurement error) obscuring the observed hormone dynamics ^3^. Several computational methods for the analysis of endocrine data have been proposed in the literature ^3,10-13^, and deconvolution analysis, is the current gold-standard method for analyzing LH pulsatility in humans ^3^. However, all methods lack an open-access, user-friendly interface that can be readily used by clinicians.

To meet these challenges, we have developed HormoneBayes, a novel, open-access Bayesian framework for the analysis of hormone pulsatility data. Unlike previous methods, HormoneBayes uses a stochastic model, describing hormone levels in the circulation incorporating measurement error, and leverages Bayesian statistics ^14^ to infer model parameters and latent variable dynamics. We show that HormoneBayes can be used to accurately identify LH pulses and estimates clinically relevant measures such as inter-pulse intervals and secretion rates. The framework also provides a handle on estimation uncertainty via Bayesian posterior distributions. We showcase how this feature can be used to enable the understanding of alterations in LH pulsatility by analyzing the effect of direct hypothalamic stimulation using the neuropeptide kisspeptin on a subject-by-subject basis. We also demonstrate that HormoneBayes can be used to analyze LH pulsatility in different clinical contexts/reproductive states (including healthy men, women before and after menopause, and women with reproductive disorders such as HA or PCOS). Importantly, the framework comes with a user-friendly, open-access graphical interface that make the core functionality of the framework easily accessible to clinicians in the clinic.

## Results

### Analyzing pulsatile hormone dynamics using the hormoneBayes framework

The hormoneBayes framework allows inference of key physiological parameters describing pulsatile hormone dynamics. The framework utilizes stochastic mathematical models describing circulating hormone levels and state-of-the-art Bayesian machinery to calibrate these models to data of hormone profiles and infer model parameters. **Figure 1** presents a parsimonious model (a simple model with great explanatory power) describing circulating LH levels. The model assumes that there are two modes of LH secretion, namely pulsatile and basal. The former corresponds to an on/off signal that randomly switches between two states (corresponding to a high and a low activity) while the latter corresponds to a continuous noisy signal. Furthermore, the model incorporates LH clearance as a linear first order process leading to an exponential decay of LH levels following a pulse ^3^. We note that more detailed clearance models, such as the bi-exponential model ^3^, could be easily integrated. By considering the processes of LH release and clearance, the model predicts LH circulating dynamics in terms of five key parameters that can be recovered from data: 1. LH clearance rate; 2. maximum LH release rate; 3. strength of the pulsatile signal relative to the basal signal; 4. mean time in the on state; 5. mean time in the off state. Moreover, the model incorporates measurement error as an additional parameter that is determined based on the assay coefficient of variation (CV).

**Figure 1.**
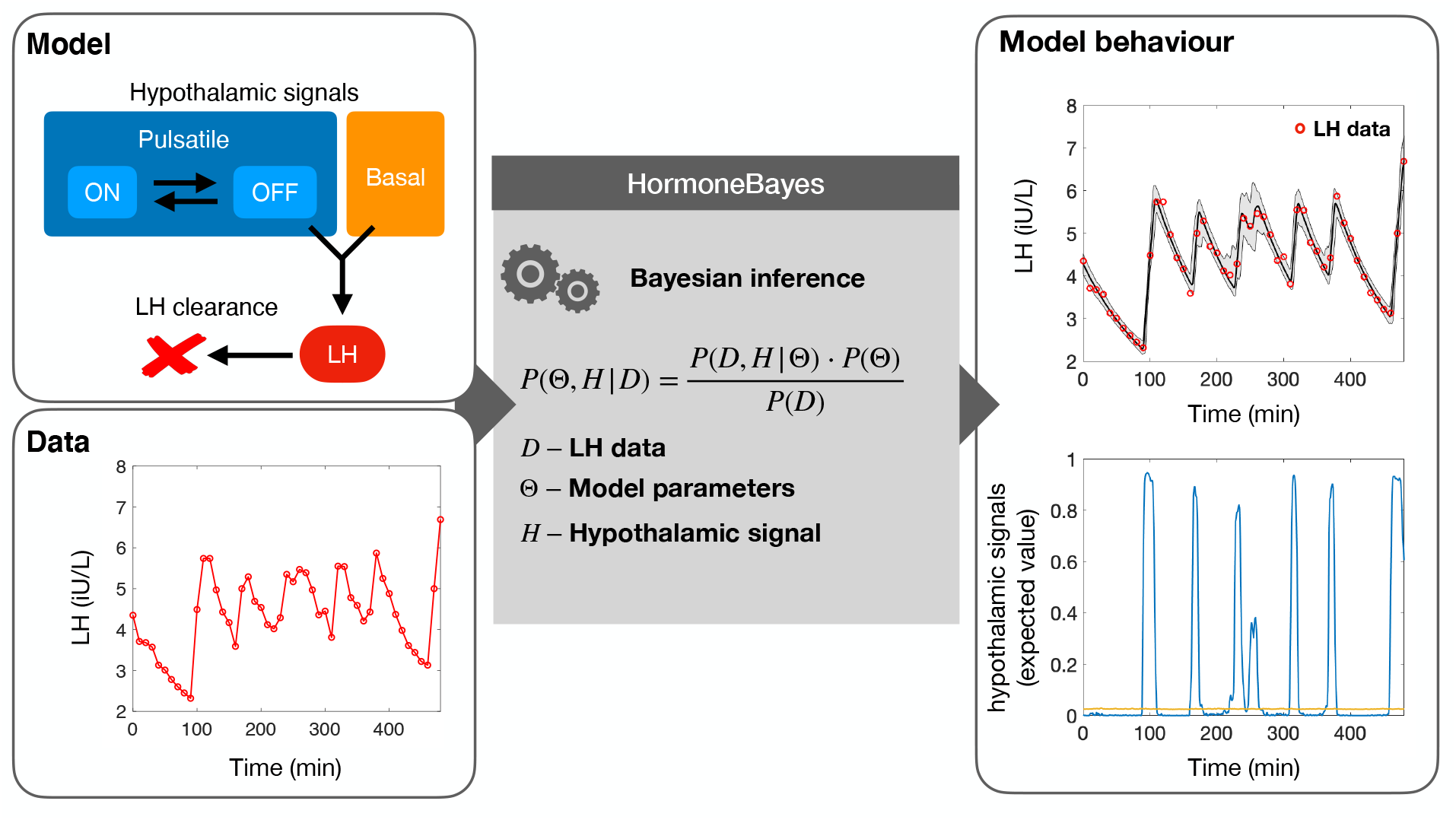
HormoneBayes: a Bayesian framework for analyzing pulsatile LH dynamics. The framework uses a parsimonious mathematical model to describe LH levels in circulation as the net effect of secretion and clearance. In the model secretion is driven by a basal hypothalamic signal and a pulsatile signal (mimicking the dynamics of the GnRH pulse generator which can be turned ‘on’ or ‘off’). An efficient Markov-Chain Monte-Carlo (MCMC) method performs the Bayesian inference and extracts model parameters and latent hypothalamic dynamics, which are compatible with the observed data.

HormoneBayes relies on the Bayesian paradigm to extract information from the observed data. Using the Bayes theorem, the method revises our prior beliefs regarding model parameters by transforming the parameters’ prior probability density distributions into posterior distributions. Parameter prior distributions enable the user to input context-specific information into the analysis, hence enhancing the flexibility of the method to handle different datasets. For example, when dealing with data from post-menopausal women the user could choose to adjust the parameter priors to acknowledge a higher LH secretion rate and/or more sustained basal secretion. **Figure 1** illustrates how the Bayesian machinery allows us to calibrate the model to the data and extract information regarding model parameter and latent hypothalamic signals with an estimate of certainty. As we explain in the sections to follow, this information can be used to identify pulses; summarise the data (e.g., providing mean and standard deviation estimates); and perform statistical tests.

### Identifying LH pulses

Using data to infer the latent hypothalamic signal provides a transparent way to identify pulses based on their likelihood under the model. As explained above, the model assumes LH pulses are partly driven by an on/off hypothalamic signal. This latent variable (i.e., inferred variable) takes two values indicating the ‘on’ (1) and ‘off’ (0) state of the hypothalamic pulse generator, and therefore the expected posterior estimate (inferred from LH profiling data) can be interpreted as the probability that at any given time the hypothalamic pulse generator is ‘on’. This quantitative measure for accessing the likelihood of a pulse can significantly ease the job a clinician trying to decide whether an upstroke in the LH profile represents a pulse or not. **Figure 2** illustrates a representative example of an LH trace with two obvious pulses occurring at times 100min and 300min. These are indeed identified by inspecting the hypothalamic signal profile which peaks at around those times. At time 200min a less pronounced bump in LH could be indicative of an LH pulse, however the inferred pulsatile hypothalamic signal remains well below 0.5, indicating that under the current model there is higher probability that the bump is a measurement artefact rather than a pulse.

**Figure 2.**
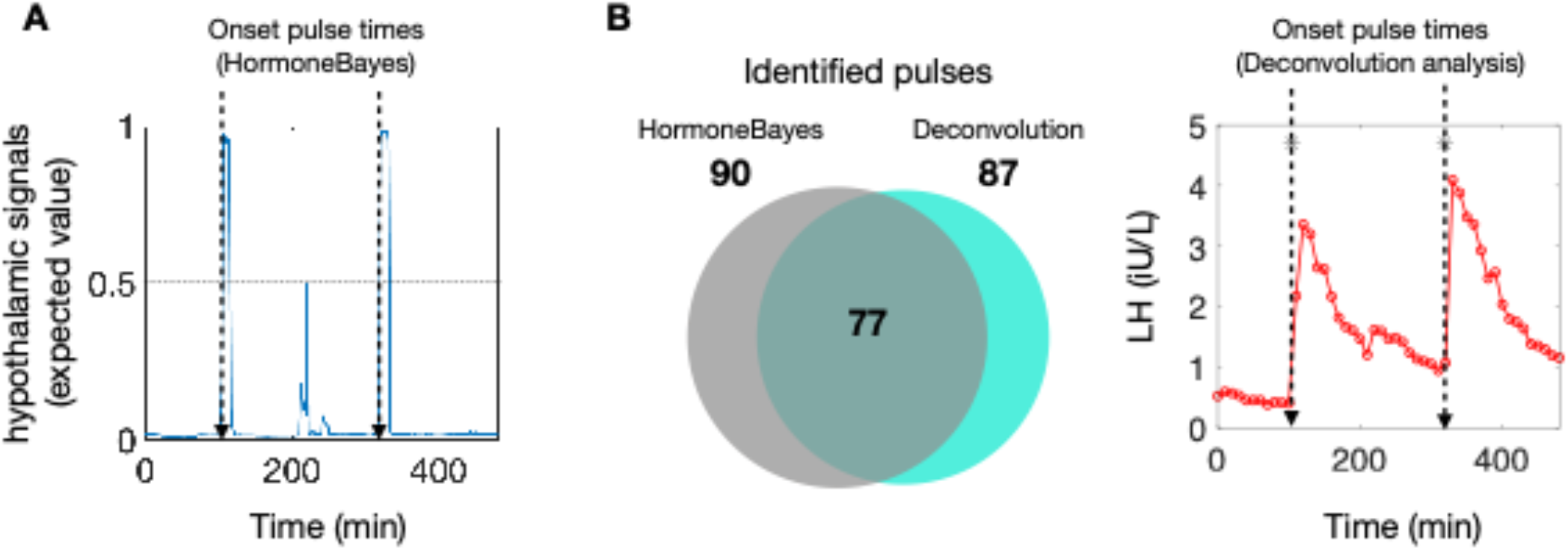
Pulse identification using HormoneBayes. **(A)** Pulses can be identified using the expected value of the pulsatile hypothalamic signal, which can be interpreted as the probability of a pulse at a given timepoint. Here, we mark the onset of a pulse when the pulsatile hypothalamic signal crosses the 0.5 threshold, indicating that at this point a pulse is the most likely event. (B) The majority of the identified pulses (89%, 77/87) are in line with those obtained using the deconvolution method. For the analysis we used LH data obtained from healthy pre-menopausal women in early follicular phase (n=16).

To validate our pulse identification method, we used a database of LH profiles obtained from healthy pre-menopausal women and compared the identified pulses with those obtained by the current gold-standard deconvolution method, which uses a maximum likelihood approach to fit a secretion model to the data ^3^. Here, we identify a pulse when the posterior probability that the hypothalamic signal is ‘on’ exceeds a threshold value. We use 0.5 as the threshold value, which signifies there is higher probability that the hypothalamic pulse generator is on (rather than off). This value yields the highest agreement between hormoneBayes and the deconvolution method (see supplementary material fig S4). We find that hormoneBayes agrees with the deconvolution method in 77 out of 87 identified pulses (89%). Moreover, 13 pulses identified by hormoneBayes were not identified by the deconvolution method. Overall, this suggests a good agreement between the two methods, with hormoneBayes having the added advantage of providing a measure for the likelihood of each pulse that clinicians and researchers can use to inform their clinical decision making.

### Variation of model parameter within and across groups

To test the applicability of hormoneBayes in different contexts we compile a database of LH profiles from four groups with diverse LH dynamics (men, healthy pre-menopausal women with regular menstrual cycles, post-menopausal women, women with HA, and women with PCOS). As illustrated in Fig. 3, the model successfully captures the differences in LH dynamics in all four groups. Moreover, the model allows us to summarize LH data through model parameters and assess the variability across and within groups. We find that two model parameters explain most of the variability between groups, namely the maximum secretion rate and pulsatility strength (**Fig 3C**). The first parameter describes how much LH can be secreted over time, whilst the second parameter quantifies the strength of the pulsatile signal relative to the basal signal. Based on these two parameters there is a strong distinction between women with PCOS and women with HA, who have lower LH secretion rates and/or diminished LH pulsatility strength (i.e., pulsatile signal is weak relative to the basal signal). Furthermore, post-menopausal women display higher secretion rates compared to pre-menopausal women but also reduced pulsatility strength (i.e., pulses are less pronounced when higher level of LH are established post menopause). Interestingly, healthy men and women illustrate a much lower parameter variability as compared to HA and post-menopausal women, which could be indicative of various degrees of severity of HA/PCOS and tighter LH regulation in healthy individuals of reproductive age.

**Figure 3.**
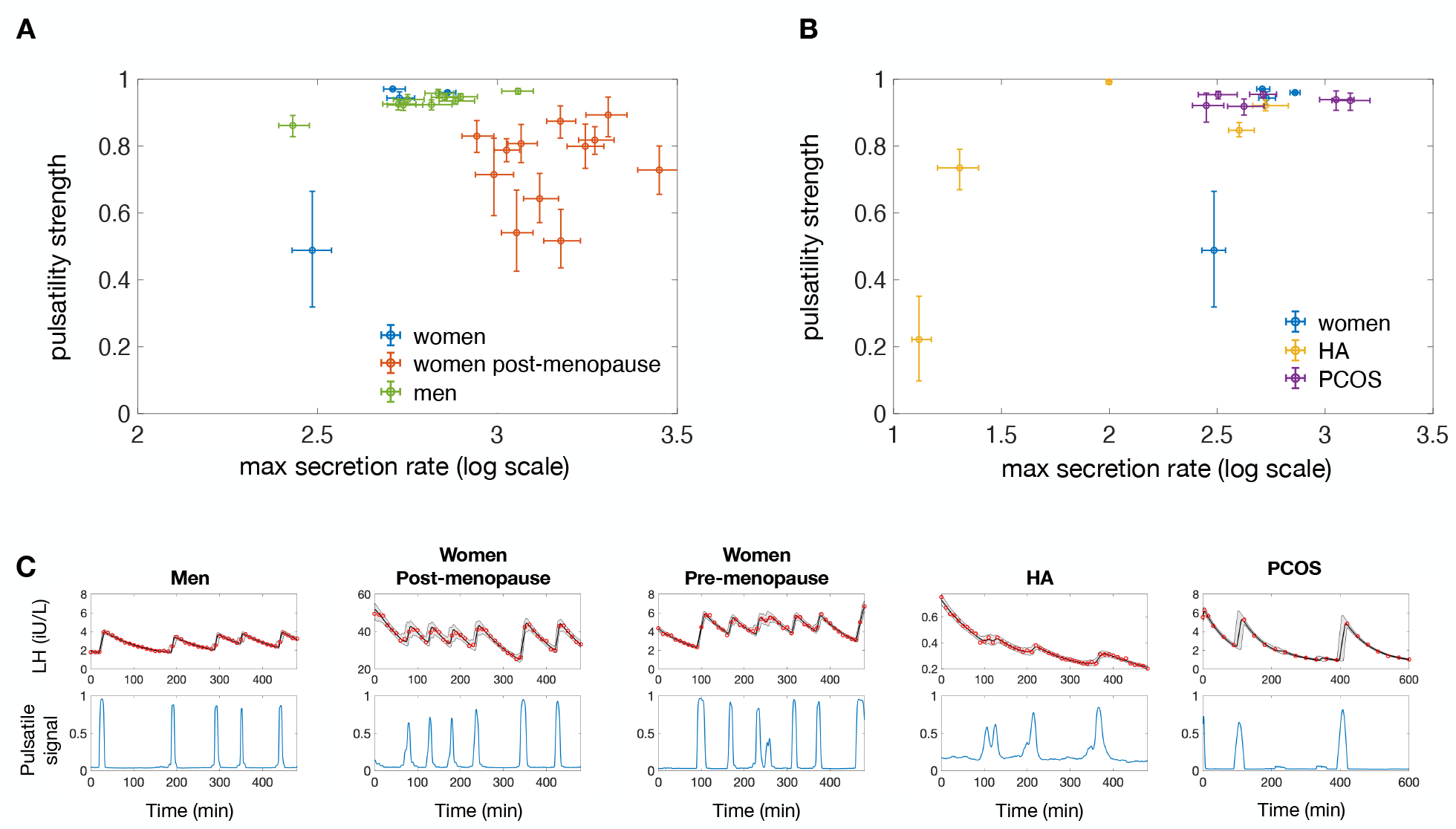
HormoneBayes handles LH pulsatility analysis in different contexts. (**A**) Inferred pulsatility strength and maximum secretion rate parameters for different individuals: healthy men (n=10); healthy post-menopause women (n=13); healthy pre-menopausal women (n=4). (**B**) Inferred parameters for healthy pre-menopausal women (n=4); women with PCOS (n=6) and women with HA (n=5) illustrating how the assessment of LH pulsatility could help facilitate diagnosis of patients presenting with reproductive endocrine disorders. (**C**) Representative fits of the model are given for one subject in each dataset.

## Discussion

We have presented HormoneBayes, a novel computational framework for analyzing hormone pulsatility. The framework combines (i) mathematical models describing hormone dynamics with (ii) computational Bayesian machinery for inferring model parameters from data. Using a parsimonious mathematical model of LH secretion, we have demonstrated the clinical utility of HormoneBayes in accurately analyzing LH pulsatility data, identifying pulses, and summarizing data in terms of model parameters to predict different clinical conditions/reproductive states on an individual patient level basis.

HormoneBayes leverages the Bayesian paradigm to infer model parameters from the data. The Bayesian approach means that hormoneBayes outputs a (posterior) density distribution for model parameters that has an intuitive interpretation, i.e, it describes the probability that parameters attain some value given the data. This enables the use of posterior distributions for statistical testing, for example to assess the effect of hormonal interventions on LH secretion parameters at the population level but also on a subject-by-subject basis. We expect such personalized analysis to become mainstream as measurement technologies mature enabling cheap sampling of hormone levels in real time ^15^.

Ultimately, data analysis using HormoneBayes is as credible as the underlying model used to describe hormonal dynamics. Here, we have used a parsimonious model that assumes two modes of LH secretion, pulsatile and basal. This assumption is in par with current physiological understanding of the system and the hypothalamic pulse generator hypothesis ^16,17^. Furthermore, to model LH circulation levels the model assumes a linear clearance rate. At least one other model used for the analysis of LH pulsatility has used more complex (multiple timescale) clearance dynamics, however we expect this assumption should have a minimal impact for the purpose of assessing LH pulsatility. Nevertheless, the modular design of HormoneBayes allows future extensions of the model with the scope of comparing how well different models capture LH dynamics as well as enabling the analysis of hormone dynamics beyond LH.

## Methods

### LH datasets

Participants included in five different clinical research studies (involving healthy men^18^, healthy pre-menopausal women^19^, post-menopausal women^20^, women with PCOS and women with HA^8^) attended our clinical research facility for a series of 8 hour study visits on separate days that included baseline (vehicle treatment) and active intervention according to their relevant trial protocol as previously described ^8,18-20^. Ethical approval for these studies was granted by: the Hammersmith and Queen Charlotte’s and Chelsea Hospitals Research Ethics Committee (registration number 05/Q0406/142)^8,19^; the UK National Research Ethics Committee-Central London (Research Ethics Committee number 14/LO/1098)^18^; and the West London Regional Ethics Committee (15/LO/1481)^20^. All studies were conducted according to Good Clinical Practice Guidelines. Once introduced to the unit, a cannula was inserted into a peripheral vein under aseptic conditions (time at least -30 minutes), through which all subsequent blood samples were taken every 10 minutes from time 0 until 480 minutes. All participants were ambulatory and could eat and drink freely during the study visit. In the kisspetin-54 study, participants received 8-hour iv infusions of vehicle and 0.1 nmol kg^-1^ hour^-1^ kisspetin-54 at different visits. All blood samples were left to clot for at least 30 minutes prior to centrifugation at 503 rcf for 10 minutes, after which the serum supernatant was extracted and immediately frozen at -20°C prior to subsequent analysis using an automated chemiluminescent immunoassay method (Abbott Diagnostics, Maidenhead, UK) in batches after study completion. Reference ranges were as follows: LH 4-14 IU/L; respective intra-assay and inter-assay coefficients of variation were 4.1% and 2.7%; analytical sensitivity was 0.5 IU/L.

### Stochastic model of LH

We used a discrete-time, stochastic model to describe pulsatile LH dynamics. The model comprises of three dynamical variables, *P*_*t*_, *B*_*t*_, and *LH*_*t*_, that describe the pulsatile and basal hypothalamic signals and the LH concertation in circulation, respectively.

The pulsatile hypothalamic signal *P*_*t*_ can take two values: *H*_*t*_ = 0 corresponding to the ON state; and *H*_*t*_ = 1 corresponding to the OFF state. The stochastic dynamics of *H*_*t*_ are governed by the following probability matrix

| | | $H_{s+\delta t}$ | |
| --- | --- | --- | --- |
|  |  | **0** | **1** |
| $H_s$ | **0** | $1 - \frac{1}{\tau_{ON}} \cdot \delta t$ | $\frac{1}{\tau_{ON}} \cdot \delta t$ |
| | **1** | $\frac{1}{\tau_{OFF}} \cdot \delta t$ | $1 - \frac{1}{\tau_{OFF}} \cdot \delta t$ |

i.e., parameters τ_*ON*_ and τ_*OFF*_ govern the probabilities that the value of *H* will either flip or remain the same over the time interval (*S, S* + *δt*).

The evolution of the basal hypothalamic signal, *B*_*t*_, is described using a discrete time autoregressive model obeying the following equation

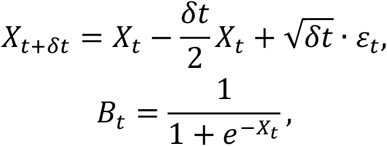

where *ε*_*t*_ is a normally distributed random variable with zero mean and unit variance. Note that both *B*_*t*_ and *H*_*t*_ are bounded in the interval [0,1].

The two hypothalamic signals drive LH secretion, and along with LH clearance dictate the circulating LH levels, *LH*_*t*_. The equation describing the time evolution of *LH*_*t*_, is

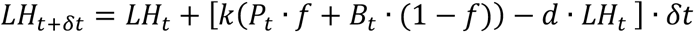

where *d* denotes the clearance rate, *k* denotes the maximum secretion rate, and parameter *f* (termed pulsatility strength) describes the relative strength of the two hypothalamic signals. Finally, the model assumes measurement error in the form:

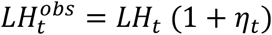

where *η*_*t*_ is a normally distributed random variable with zero mean and std. deviation equal to the CV of the assay. Throughout our analysis we have used *δt* = 1 min, hence, assuming that the system dynamics do not change significantly over shorter times.

··L^-1^

### Bayesian inference

The hormoneBayes framework uses Bayesian inference to obtain model parameters Θ = (τ_*ON*_, τ_*OFF*_, *k, d, f*) and latent variable (*H*_*t*_, *B*_*t*_) dynamics from LH profiling data *D*. In particular, hormoneBayes solves the inference problem by sampling from the target posterior distribution:

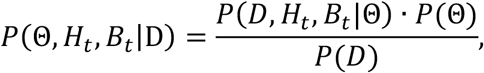

where *P*(Θ) is the prior parameter distribution; *P*(*D, H*_*t*_, *B*_*t*_|Θ) = *P*(*D*|Θ, *H*_*t*_, *B*_*t*_) × *P*(*H*_*t*_, *B*_*t*_) is the likelihood of the data given the parameters; and *P*(*D*) = ∫ *P*(*D, H*_*t*_, *B*_*t*_|Θ) × *P*(Θ) is the marginal likelihood or model evidence.

Sampling from the full posterior distribution is performed using a Gibbs sampler, which is an iterative Monte Carlo Markov Chain (MCMC) scheme. The algorithm is initialised with parameter values drawn from the prior distribution, i.e., Θ^0^∼*P*(Θ) and each subsequent iteration, *i* = 1, …, *M* involves two steps: (1) sampling latent variables (*H*_*t*_, *B*_*t*_)^*i*^ given the data, D, and the current parameter values Θ^i−1^ and (2) sampling new parameter values Θ^*i*^ given the D and the latent variables (*H*_*t*_, *B*_*t*_)^*i*^. The first step is performed using Sequential Monte Carlo (SMC) with ancestral sampling. The second step is further broken down into two parts, first parameters (τ_*ON*_, τ_*OFF*_) are sampled using an adaptive Metropolis-Hastings sampler and then parameters (*k, d, f*) are sampled using Hamiltonian Monte Carlo (HMC).

To access the effect of pharmacological interventions on LH pulsatility, hormoneBayes allows the input of two LH profiling datasets, corresponding to control and perturbed conditions. The inference task is then modified to allow inference of parameters Θ_*c*_ = (τ_*ON*_, τ_*OFF*_, *k, d, f*) for the control case as well of four additional parameters Θ_*P*_ = (*p*τ_*ON*_, *p*τ_*OFF*_, *pk, pf*) corresponding to the perturbed case. In mathematical terms the target posterior is now given by

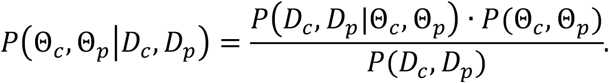

Sampling from the posterior is performed as described above. We quantify the effect of the perturbation as the log-ratio of perturbed to control values.

Independent prior distributions were used for all model parameters throughout this study: log_10_τ_*ON*_∼*U*(*log*10(5), *log*_10_(240)) and log_10_τ_*OFF*_∼*U*(*log*10(5), *log*10(240)), based on the sampling rate (10min) and duration (480min) used in the LH profiling studies; log_10_(*k*)∼*N*(0,5), set as a broad uninformative prior; 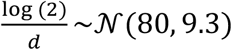, based on LH half-life data; and log(*f*)∼ *U*(0,1), to accommodate analysis of different LH profiles with high or low pulsatility. Evaluation of the algorithm on synthetic dataset can be found in supplementary material fig S1-3. An open access implementation of the algorithm along with a supporting graphical interface can be downloaded from https://git.exeter.ac.uk/mv286/hormonebayes.

## Data Availability

All data produced in the present study are available upon reasonable request to the authors

## Acknowledgements

MV and KTA acknowledge the financial support of the EPSRC via grants EP/T017856/1 and EP/N014391/1, and BBSRC via grants BB/S000550/1 and BB/S001255/1. JKP is supported by a NIHR academic fellowship, MRC (MR/M024954/1), and Expanding Excellence in England (E3) - Exeter Diabetes Research Unit. AA is supported by an NIHR Clinician Scientist Award (CS-2018-18-ST2-002). WSD is supported by an NIHR Senior Investigator Award.

