## Supplemental material for "HormoneBayes: a novel Bayesian framework for the analysis of pulsatile hormone dynamics"

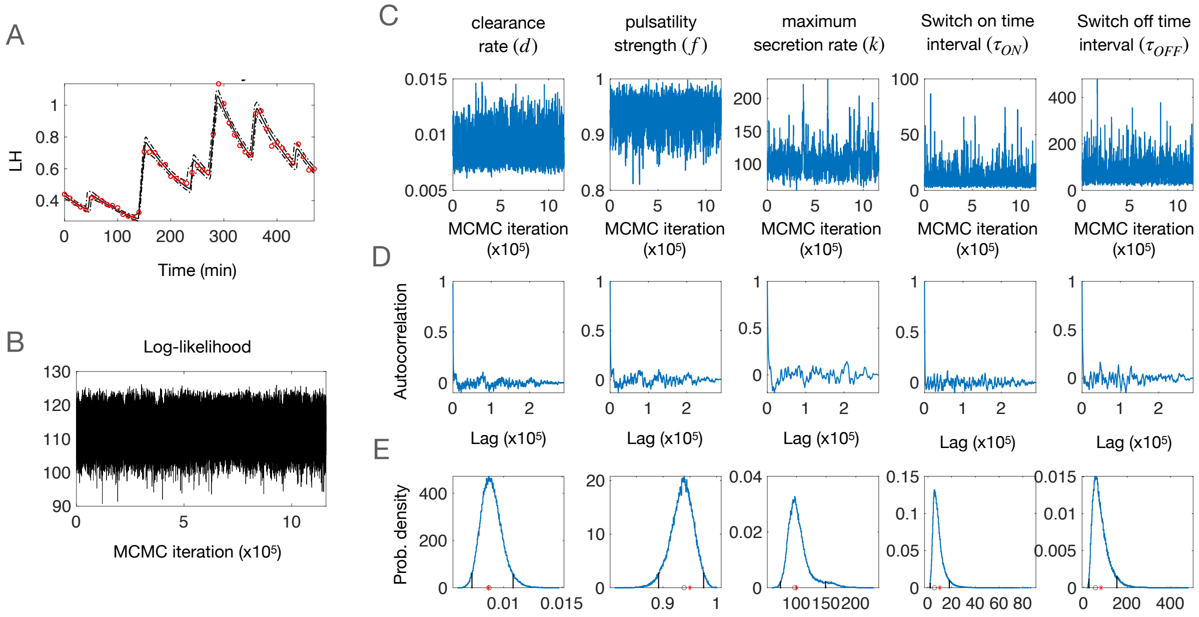


**Figure S1.** We tested HormoneBayes by fitting the model to synthetic data. (A) Representative example of synthetic data (red circles) along with the model fit generated by HormoneBayes. In this example, synthetic data were generated using the model described in the main text with the following parameter values: $d = 0.0087$, $f = 0.95$, $k = 100$, $\tau_{ON} = 10$, $\tau_{OFF} = 80$. Four independent MCMC chains were generated using the algorithm described in the main text, with each chain consisting of 3 $10^5$ iterations (the first $10^4$ iterations were excluded from further analysis). Traces of the (B) log-likelihood and (C) model parameters. Convergence of the MCMC chains was assessed using their autocorrelation functions (D) and the potential scale reduction factor (R-hat statistic), which in all cases was between 1 and 1.005 indicating good convergence. (E) The posterior parameter distributions accurately reflect the actual parameter values (black asterisks). In each figure, the red circle denotes the maximum a posteriori (MAP) estimate and the vertical black lines the Highest Posterior Density (HPD) credible interval.

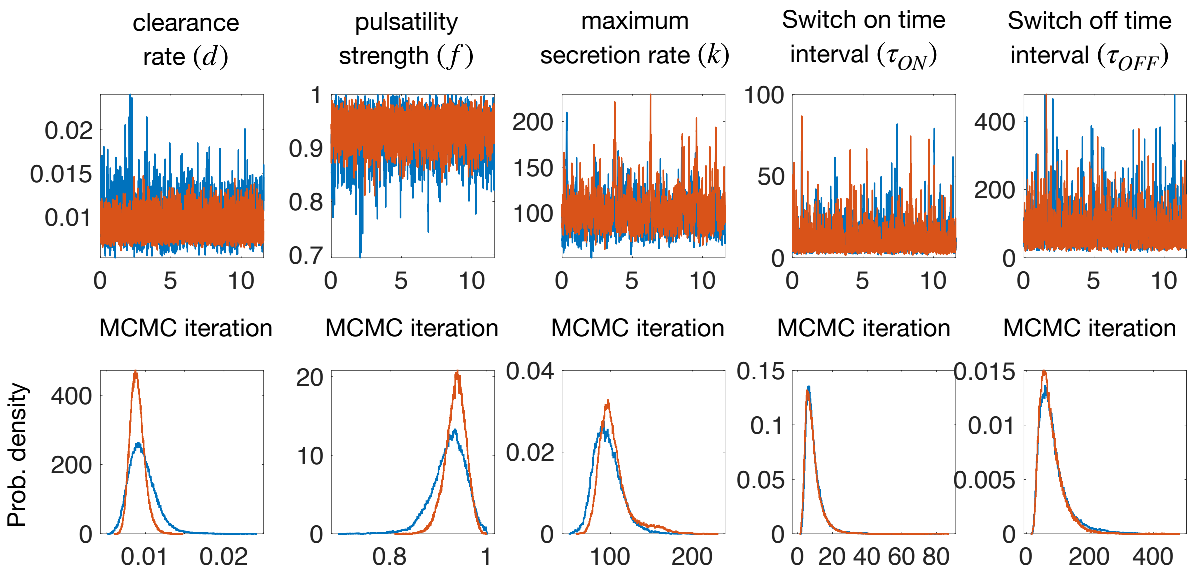


**Figure S2.** We used HormoneBayes to analyse synthetic LH data under two different specifications of the prior for the LH clearance parameter. In the first case (blue) we used an informative prior ( $\log(2) \cdot d^{-1} \sim \mathcal{N}(80{,}9.3)$ ). In the second case (red) we used an uninformative prior (uniform $\log(2) \cdot d^{-1} \sim \mathcal{N}(10^{-5}, 10^{5})$). In both cases the MCMC chains converged to similar posterior distributions that reflect the parameters used to generate the data. Synthetic data were generated using the model presented in the main text with the following parameters: $d = 0.0087$, $f = 0.95$, $k = 100$, $\tau_{ON} = 10$, $\tau_{OFF} = 80$.

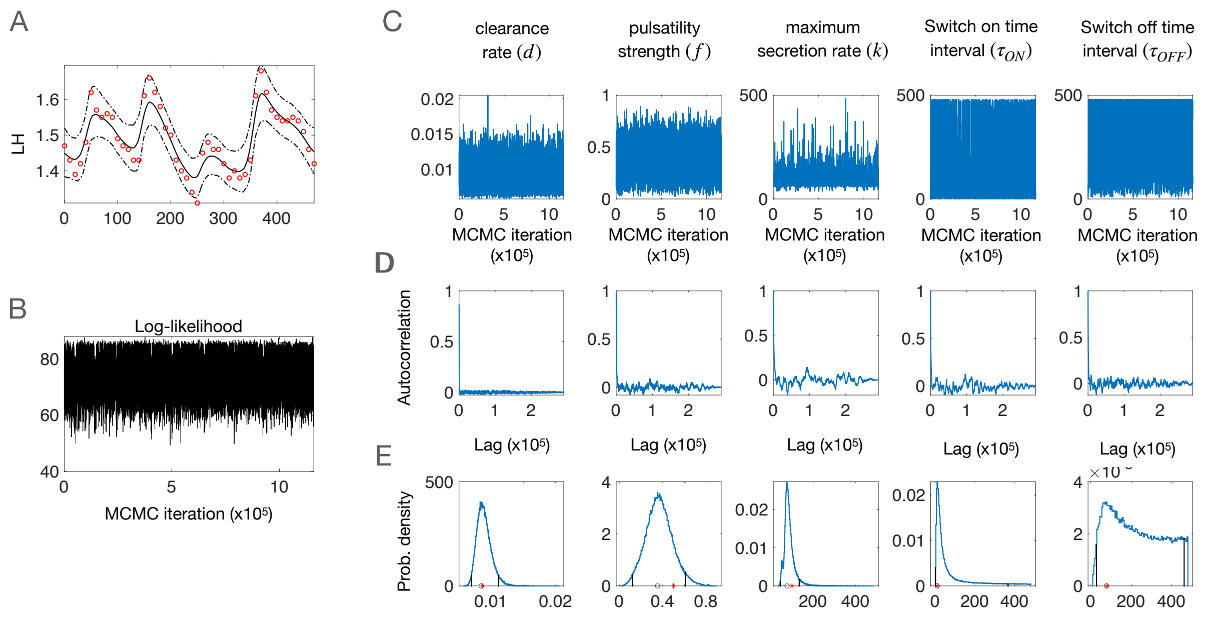


**Figure S3.** When pulses are not clearly evident in the data a more informative prior on parameter f can used. (A) Representative example of synthetic data (red circles) along with the model fit generated by HormoneBayes. In this example, synthetic data were generated using the model described in the main text with the following parameter values: $d = 0.0087$, $f = 0.5$, $k = 100$, $\tau_{ON} = 10$, $\tau_{OFF} = 80$. Here, a Beta distribution with parameters $\alpha = 4$, $\beta = 6$ was used as a prior for parameter $f$ instead of a uniform. Four independent MCMC chains were generated using the algorithm described in the main text, with each chain consisting of 3 $10^5$ iterations (the first $10^4$ iterations were excluded from further analysis). Traces of the (B) log-likelihood and (C) model parameters. Convergence of the MCMC chains was assessed using their autocorrelation functions (D) and the potential scale reduction factor (R-hat statistic), which in all cases was found to be between 1 and 1.009 indicating good convergence. (E) The posterior parameter distributions provide some information about the actual parameter values (black asterisks). In each figure, the red circle denotes the maximum a posteriori (MAP) estimate and the vertical black lines the Highest Posterior Density (HPD) credible interval.

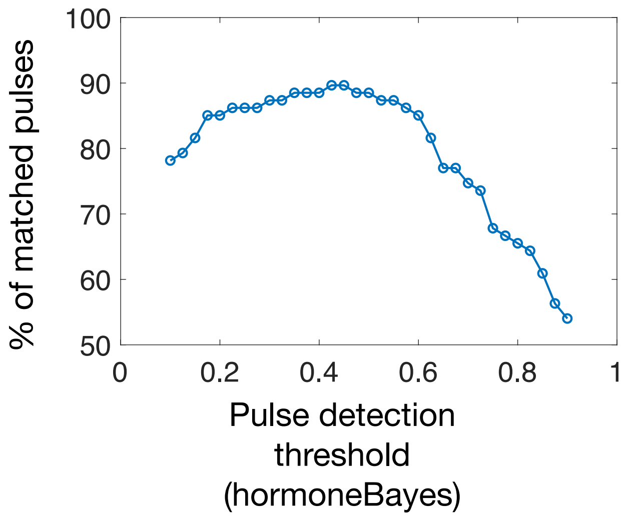


**Figure S4.** Pulse identification using HormoneBayes. Percentage of matched pulses between hormoneBayes and the Deconvolution method as the pulse-detection threshold is varied. Maximum agreement between the two methods is observed when the threshold is around the 0.5 range. For the analysis we used LH data obtained from healthy pre-menopausal women in early follicular phase (n=16).
